# Data and care integration for post-acute intensive care program of stroke patients: effectiveness assessment using a disease-matched comparator cohort

**DOI:** 10.1101/2022.04.13.22273573

**Authors:** Emili Vela, Aina Plaza, Gerard Carot-Sans, Joan C. Contel, Mercè Salvat-Plana, Marta Fabà, Andrea Giralt, Aida Ribera, Sebastià Santaeugènia, Jordi Piera-Jiménez, the REDOM group

**Affiliations:** System Information Area, Catalan Health Service, Barcelona, Spain; Digitalization for the Sustainability of the Healthcare system (DS3), IDIBELL, Barcelona, Spain; Ministry of Health, Catalan Government, Barcelona, Spain; Chronic Care Program. Department of Health. Barcelona, Spain; Central Catalonia Chronicity Research Group (C3RG), Centre for Health and Social Care Research (CESS), University of Vic-Central University of Catalonia (UVIC-UCC), Vic, Spain; Catalan Stroke Program, Health Department, Agency for Health Quality and Assessment (AQuAS), CIBER Epidemiología y Salud Pública. Barcelona, Spain; Direcció d’Estratègia i Nous Projectes, Institut Municipal de Serveis Socials, Ajuntament de Barcelona, Barcelona, Spain; Institut Municipal de Serveis Socials, Ajuntament de Barcelona, Barcelona, Spain; Cardiovascular Epidemiology and Outcomes Research Unit. Vall d’Hebron Institut de Recerca (VHIR), CIBER Epidemiología y Salud Pública. Barcelona, Spain; Programa de Prevenció i Atenció a la Cronicitat i Pla Director Sociosanitari, Departament de Salut. Programa d’Atenció Integrada Social i Sanitària (PAISS), Departament de Salut, Barcelona, Spain; Facutly of Informatics, Telecomunications and Multimedia, Universitat Oberta de Catalunya, Barcelona, Spain; Ajuntament de Barcelona; Institut Català de la Salut; Hospital Clínic; Hospital de Sant Pau; Parc de Salut Mar; Mutuam Güell; Parc Sanitari Pere Virgili; Fisiogestión; Departament de Salut

**Author notes:** **Corresponding author** Jordi Piera-Jiménez, Catalan Health Service.

## Abstract

**Purpose:** To assess the effectiveness of an integrated care program for post-acute care of stroke patients, the return home program (RHP program), deployed in Barcelona (North-East Spain) between 2016 and 2017 in a context of health and social care information systems integration.

**Design:** The health outcomes and resource use of the RHP program participants were compared with a population-based matched control group built from central healthcare records of routine care data.

**Findings:** The study included 92 stroke patients attended within the RHP program and their matched-controls. Patients in the intervention group received domiciliary care service, home rehabilitation, and telecare significantly earlier than the matched-controls. Within the first two years after the stroke episode, recipients of the RHP program were less frequently institutionalized in a long-term care facility (5% vs. 15%). The use of primary care services, non-emergency transport, and telecare services were more frequent in the RHP group.

**Originality:** Our analysis shows that an integrated care program can effectively promote and accelerate delivery of key domiciliary care services, reducing institutionalization of stroke patients in the mid-term. The integration of health and social care information allows not only a better coordination among professionals but also to monitor health and resource use outcomes of care delivery

## Introduction

The continuity of care during transitions between different places or settings is the cornerstone of patient-centered integrated care. Of all these transitions, hospital discharge after a disabling health event has remarkable impact on patients’ life, particularly those with continuous complex care needs [1]. The difficulties associated with discharge of these patients can be overcome by appropriate comprehensive discharge plans that consider education on self-management, coordination of care providers, medication reconciliation, and assessment of financial barriers, among others [1,2]. Comprehensive discharge plans have shown to reduce post-discharge mortality, emergency department visits, and readmissions in patients with chronic complex diseases transiting from hospital to primary care [2].

Stroke affects over 80 million people worldwide, and it is leading cause of disability among adults [3]. Owing to their limited capacity for activities of daily living, stroke patients often require domiciliary care after hospital discharge [4]. Home care services, aimed to support people in their activities of daily living, are often prescribed and/or provided by social care services. Hence, the continuity of care in this transition strongly depends on the adequate integration between social and health care services that identifies the patient needs from all spheres of care. Unsuccessful integration often results in a delay of domiciliary rehabilitation and care, which may worsen the health outcomes in stroke patients [5–7].

The healthcare burden of stroke is expected to increase with the population shift forecasted for the upcoming years. Although projections estimate a slight decrease of disability-adjusted life years despite the increasing incidence [8], the raising number of people with rehabilitation needs is likely to impact health and social care systems in high-income countries. Furthermore, the aging population will increase the proportion of patients with continuous complex care needs, including domiciliary care services, contributing to the overload of healthcare services. This scenario stresses the need for implementing integrated care pathways that efficiently screen stroke patients and design adequate post-discharge care plans that cover both the social and health care needs of patients. In this study we assessed the health outcomes and resource use of a post-stroke intensive home care program based on the integration of social and health care for improving domiciliary care of stroke patients after hospital discharge.

## Methods

### Study Design and Patients

This was a retrospective, matched-control study to assess the effectiveness of an integrated care program for post-acute care of stroke patients (i.e., the return home program [RHP]) in Barcelona (North-East Spain). The study included all consecutive patients entering the program between February 15, 2016 and February 15, 2017. All patients with domiciliary care needs were consecutively screened for the program. Inclusion criteria were living alone or with another person with limited capacity to deliver adequate care, previous dependency, cognitive impairment, and stroke severity (NIHSS >10)).

A comparator group was built using patients from the general population admitted to any of the tertiary hospitals of Barcelona (Hospital Clínic, Hospital del Mar, Hospital de Sant Pau, and Hospital Vall d’Hebron) within the same period. Patients living in a long-term care facility, with missing data in any of the matching variables, and those who died during hospital stay or within the 6 months following the stroke episode were not considered for the control group. Case-control matching was performed at 1:4 ratio based on sex, age (±5 years), and their healthcare risk, estimated using the morbidity adjusted groups (GMA). Briefly, the GMA is a population-based case-mix tool that allows stratifying the general population into mutually exclusive health-risk groups based on all chronic conditions and recent acute diagnostic codes [Atón Primaria]. The tool has shown high predictive capacity of key clinical and resource utilization outcomes, including mortality, hospitalizations, and visits to primary care, polypharmacy, and overall expenditure [9,10]. The selection of control individuals was prioritized according to the matching of the following conditions: type of stroke (bleeding stroke, ischemic stroke, and others), annual income (grouped into four categories of pharmaceutical co-payment), admission to intermediate care after acute care discharge, place of residence, and least age difference.

#### Intervention and Usual care

The RHP was developed to improve the post-stroke rehabilitation process and promoting the achievement of the highest rate of autonomy in the shortest possible time. The specific objectives of the RHP were (1) to identify stroke patients with social care needs during hospital stay, (2) to prescript and activate the appropriate domiciliary care according to the needs of the patient and his/her environment before hospital discharge, and (3) to coordinate synchronous interventions of rehabilitation, social care, and primary care. The RHP intervention includes three consecutive phases: candidate screening, assessment and prescription of social needs, and activation of domiciliary care provided by the local social care service before patient discharge (Figure S1, Supplementary appendix). Candidate screening occurred within the 48 hours following hospital admission; candidates were considered to have social needs if they met at least one the following indicators: living alone or with another person with limited capacity to deliver adequate care, previous dependency, cognitive impairment, and stroke severity (NIHSS >10). Once the patient was considered suitable for the RHP, the hospital-based social worker prescribes the social care resources needed for a 8-week period. The main social services prescribed at home were individual care supporting for activities of daily living, house cleaning, telemedicine, meals-on-wheels, and rehabilitation aids. Patients and their families were provided with the planned date for discharge; 72 hours before discharge from acute hospital (and 5 days before discharge from intermediate care center) the prescribed services were notified to the city Social Care Services and activated.

Patients receiving usual care were assessed by the hospital-based social worker for dependency based on their clinical situation and social environment. In case the social worker considered that the patient required domiciliary care, two separate requests were made to the primary care team (managed by the autonomous government) and to the social care team (managed by the city council). The city social care team assessed the patients again and included them in a general waiting list and contacted them when possible to start a domiciliary care program.

#### Information System Integration

Since year 2007, all centers in our area share the health-related information (i.e., clinical course, test results, digital imaging, etc.) through the Shared Electronic Health Record of Catalonia (“Història Clínica Compartica de Catalunya” [HC3]) with the main aim to improve the continuity of care [11]. Since 2015, an exchange of health and social care information was agreed among the public Catalan health system and the Barcelona City Council [12]. This exchange includes information regarding the social context, grade and characteristics of dependency, home care resources, pharmacy, discharge planning, and a summary of the individual healthcare plan. The HC3 can be accessed by the hospital social worker and the physicians and nursing teams assigned to the patient. On the other hand, information regarding social care needs and resources is collected and handled by the Barcelona City Council. Social data can be accessed by the city council social worker. Furthermore, healthcare data are periodically transferred to the Ministry of Health of the autonomous community of Catalonia for healthcare planning and analytical purposes, including public health and resource utilization.

For the RHP project, we created an ad hoc registry of stroke patients entering the program that gathered relevant health and social information of the patient. The two entities governing the datasets (i.e., the Catalonia ministry of health and the Barcelona City Council) signed an agreement for data transfer, which defined the governance of the integrated registry, the access profile, and measures for ensuring confidentiality and adherence to the organic low 15/1999 on personal data protection.

### Study Outcomes and Data Sources

The primary objective of the study was to assess the time to key events, including adverse endpoints (i.e., death and institutionalization in a long-term care facility) and service provision endpoints (i.e., receiving domiciliary care, telecare, and at-home rehabilitation services). Other outcomes included health and social care resource utilization. Health care expenditure was estimated as described before [13], whereas social expenditure was provided by the Social Service Institute of Barcelona. Resource utilization included the monthly number or average of total hospital admissions, emergency hospital admissions, visits to the emergency department, visits to a specialist, drug units (packages) dispensed by the community pharmacist, non-emergency medical transport rides, domiciliary rehabilitation visits, and contacts with primary care services, total and grouped according to the following services: general practitioner, nurse, social worker, domiciliary care team, and remote consultations.

### Statistical Analysis

Quantitative variables were described using the mean and median, whereas categorical variables were described as the frequency and percentage over available data. Time-to-event analyses were performed using the Kaplan-Meier estimate and compared with the log-rank test. Categorical variables were compared using the chi-squared test and continuous variables with the T Student’s test, except expenditure, which was not normally distributed and was compared using the Kruskal-Wallis test. The between-group differences in resource utilization and expenditure were assessed using a ratio of monthly mean utilization rates or expenditure and computed using generalized linear mixed models (Poisson for resource utilization and lognormal for expenditure), considering the random effects for matched patients. The models were adjusted for age, sex, annual income, GMA status, type of stroke, admission to intermediate care center after acute care discharge and healthcare expenditure during the year preceding the stroke episode. The significance threshold for all analyses was set at a 5% two-sided alpha error. All analyses were conducted with R [14].

## Results

### Study Participants

During the study period, 92 patients entered the RHP program after an ischemic (n=71; 77.2%) or a hemorrhagic (n= 18; 19.6%) stroke. Table 1 summarizes the main demographic, clinical, and resource use characteristics of study patients and the matched population-based control group. The two groups were similar in all characteristics, except for type of stroke, which was more dominantly ischemic in the control group. The previous healthcare resource utilization, not included among the pairing criteria, was also similar in the two groups.

**Table 1:** Characteristics of the study population and matched comparator group (Controls) at baseline.

|  | Global | Casos | Controls | p |
| --- | --- | --- | --- | --- |
|  | N=460 | N=92 | N=368 |  |
| Sexe: |  |  |  |  |
| Homes | 215 (46.7%) | 43 (46.7%) | 172 (46.7%) | 1.000 |
| Dones | 245 (53.3%) | 49 (53.3%) | 196 (53.3%) |  |
| Edat mitjana (DS) | 77.2 (9.56) | 77.0 (9.75) | 77.3 (9.52) | 0.764 |
| Trams de renda: |  |  |  |  |
| Mitjana i alta (>18.000€) | 140 (30.4%) | 27 (29.3%) | 113 (30.7%) | 0.266 |
| Baixa (<18.000 €) | 307 (66.7%) | 60 (65.2%) | 247 (67.1%) |  |
| Molt baixa (rendes socials) | 13 (2.83%) | 5 (5.43%) | 8 (2.17%) |  |
| Morbiditat prèvia a l'alta |  |  |  |  |
| Càrrega de morbiditat (Pes GMA) | 19.9 (13.6) | 19.9 (13.1) | 19.9 (13.8) | 0.998 |
| Quintils de GMA: |  |  |  |  |
| Risc molt baix | 80 (17.4%) | 16 (17.4%) | 64 (17.4%) | 1.000 |
| Risc baix | 145 (31.5%) | 29 (31.5%) | 116 (31.5%) |  |
| Risc moderat | 100 (21.7%) | 20 (21.7%) | 80 (21.7%) |  |
| Risc alt | 55 (12.0%) | 11 (12.0%) | 44 (12.0%) |  |
| Risc molt alt | 80 (17.4%) | 16 (17.4%) | 64 (17.4%) |  |
| Tipus d'ictus |  |  |  |  |
| Hemorràgic | 82 (17.8%) | 18 (19.6%) | 64 (17.4%) | 0.009 |
| Isquèmic | 375 (81.5%) | 71 (77.2%) | 304 (82.6%) |  |
| Altres | 3 (0.65%) | 3 (3.26%) | 0 (0.00%) |  |
| Ingrés en un centre sociosanitari immediatament postalta |  |  |  |  |
| No | 293 (63.7%) | 50 (54.3%) | 243 (66.0%) | 0.050 |
| Si | 167 (36.3%) | 42 (45.7%) | 125 (34.0%) |  |
| Despesa durant l'any previ a l'ictus (mitjana i [mediana]) |  |  |  |  |
| Serveis sanitaris ambulatoris | 915 [674] | 884 [692] | 923 [667] | 0.899 |
| Serveis sanitaris d'internament | 1.633 [0] | 1.537 [0] | 1.657 [0] | 0.130 |
| Farmàcia | 908 [385] | 630 [278] | 977 [396] | 0.072 |
| Altres serveis sanitaris | 121 [0] | 334 [0] | 68 [0] | 0.978 |
| Total despesa sanitària | 3.576 [1.508] | 3.385 [1.497] | 3.624 [1.508] | 0.943 |

### Clinical and Healthcare Outcomes

The survival analysis did not reveal significant differences between cases and controls regarding mortality after a stroke event (Figure 1A). On the other hand, patients in the control group were more likely to be institutionalized earlier than those in the intervention group (Figure 1B); two years after the stroke episode, the proportion of patients institutionalized in a long-term care facility was 14.2% (n=51) and 4.8% (n=4) in the control and intervention groups, respectively.

**Figure 1.**
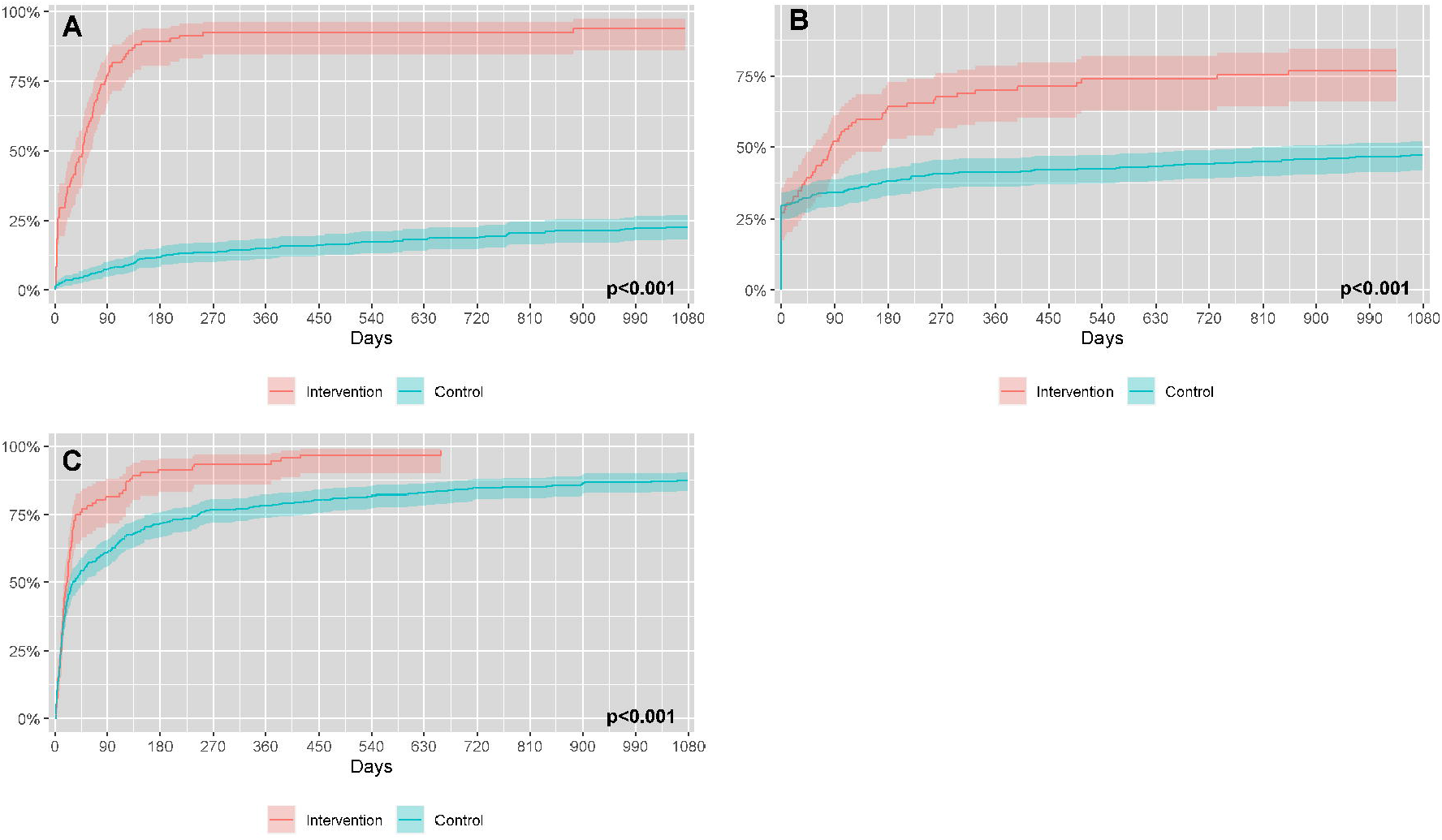
Survival analysis of relevant health and healthcare outcomes after the index stroke episode. **A:** all-cause mortality. **B:** Institutionalization in a long-term care facility.

Patients in the intervention group were more likely to receive earlier key social and health home care services (Figure 2). Two years after the index stroke episode, domiciliary care was being provided to 85 (92.4%) of patients in the RHP group and 69 (19.1%) in the control group (Figure 2A). Likewise, telecare services, which were provided to nearly one third of individuals before the stroke episode (27.2% and 29.6% in the RHP and control groups, respectively), amounted to 75.5% (n=67) in the RHP group and 44.5% (n=163) in the control group two years after the index episode (Figure 2B). At-home rehabilitation services were provided early in the two groups. However, a greater percentage benefited from this service in the RHP group; two years after the index stroke episode, the proportion of patients receiving at-home rehabilitation services amounted to 90 (98.4%) and 311 (84.8%) in the RHP and control group, respectively (Figure 2C).

**Figure 2.**
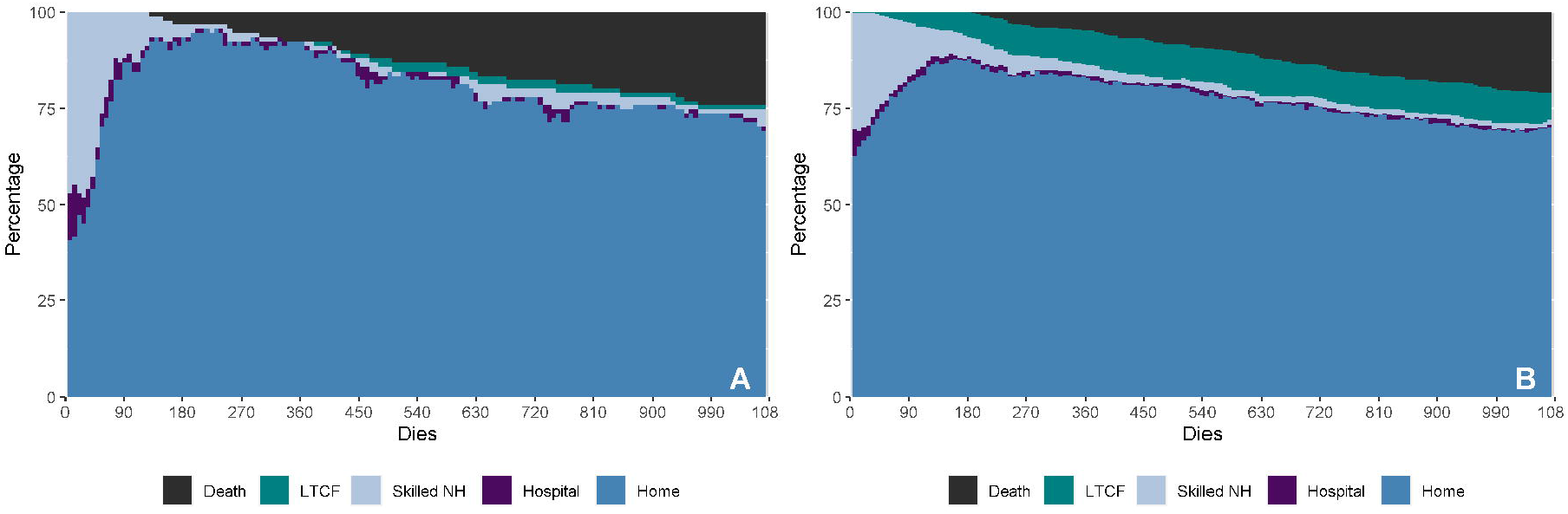
Health and social care provision after the index stroke episode. A: domiciliary care

Figure 3 summarizes the place of stay of stroke patients within the two years following the index episode. The proportion of patients at home in the RHP increased rapidly and remained higher than in the control group until the end of the follow-up, when the proportion of patients at home in the RHP and control group were 92.3% and 83.4%, respectively. Patients in the control group stayed more frequently in a long-term care facility throughout the follow-up period.

**Figure 3.**
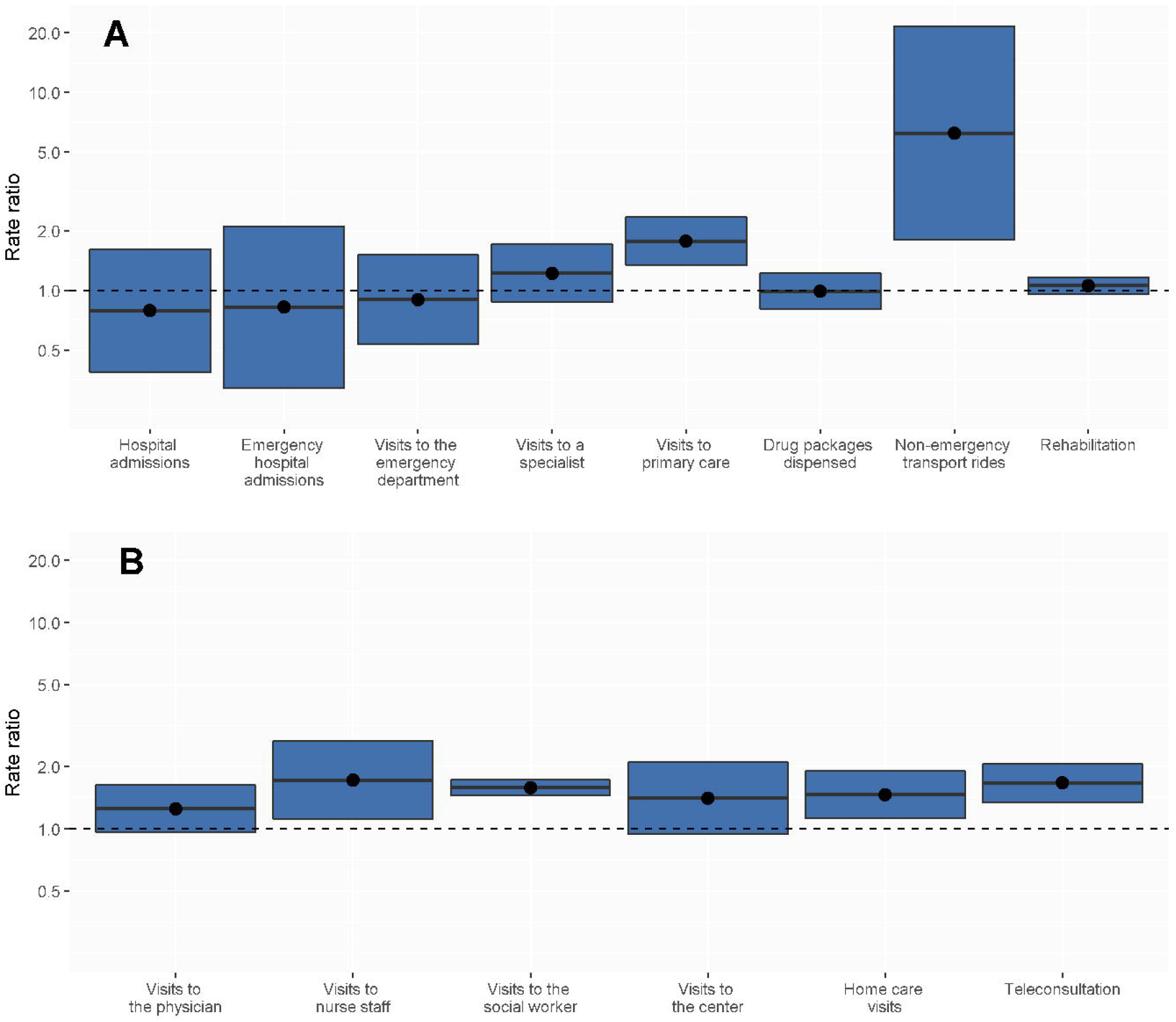
Place of stay of patients in the control (A) and intervention (B) groups within the two years following discharge from the index episode.

### Resource utilization

Within the first month following the index stroke episode, the mean social and health care expenditure was remarkably higher in the RHP group (€ 3,152) than the control group (€ 2,364), mostly due to the higher proportion of patients institutionalized in an intermediate care facility immediately after the stroke event. The monthly mean expenditure attributed to social services, intermediate care facility, outpatient visits, emergency visits, hospitalization, pharmacy, and primary care within the first year after the stroke episode is summarized in Figure S1 (Supplementary appendix). Between months 2 and 12 after the index episode, the mean monthly expenditure of the RHP and the control group were€ 784.58 and€ 657.15, respectively. The between-group differences in expenditure were more notable in the social care (€ 257.95 vs.€ 156.40) than the health care provision (€ 526.64 vs.€ 500.75). The mean monthly expenditure for the 1-to-12 moths and 2-to-12 months period according to the type of resource is provided in Table S1.

Figure 4 summarizes the difference in use of social and health care services between patients in the control and intervention group within the year following discharge (generalized mixed models); the monthly use of each resource within the two years following discharge is shown in Figure S2. The utilization of most services did not change significantly between groups. However, the overall expenditure was significantly higher in the intervention group, mostly due to the higher expenditure in social services and ―less pervasive― the number of visits to primary care (Figure 4A). Of all primary care services, non-emergency transport, and telecare services were those which increased more in the intervention group (Figure 4B).

**Figure 4.**
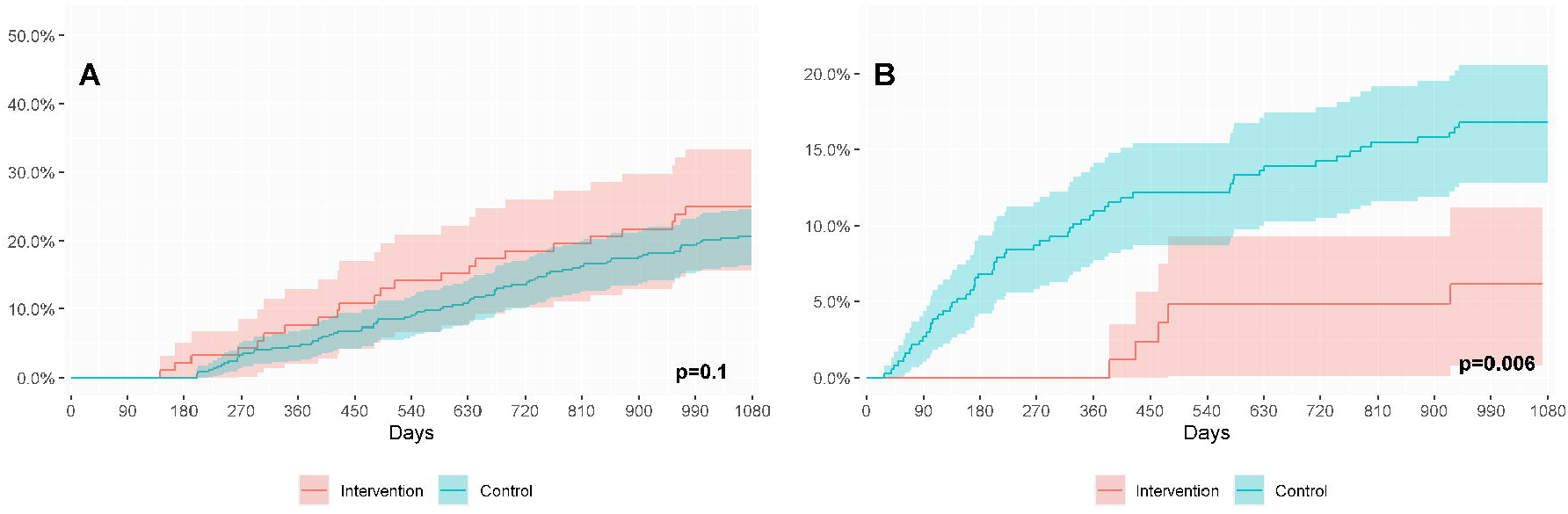
Difference in resource utilization between the intervention and control groups within the year following discharge. **A:** All services. **B:** Primary care services. The central line represent the monthly rate, adjusted using a generalized linear model; values >1 indicate higher average in the intervention group. The limits of the box indicate the 95% confidence interval.

## Discussion

Our retrospective analysis of an integrated care program for stroke patients discharged from the hospital showed that program recipients received key rehabilitation services, such as intensive post-acute home care and support for activities of daily living, earlier than stroke patients managed as usual. Of note, most stroke patients in the population-based matched control group eventually received domiciliary care services. Considering that, in the absence post-discharge supporting services (including adequate rehabilitation), most patients with stroke experience a progressive decline in functional mobility soon after discharge [7,15], this finding suggests that the later start of service provision among control patients was due to delay in planning, rather than the lack of need.

A remarkable finding of our analysis was that recipients of the RHP program were admitted in a long-term care facility less frequently and later. Thus, although stroke patients have higher risk for admission to long-term care facilities [16,17], this finding suggests that institutionalization is sometimes a consequence of inadequate or late domiciliary care provision. The dynamics of institutionalization in nursing homes have a remarkable impact on patients’ life, but also on the sustainability of the system. The implementation of a program for promoting domiciliary care is expected to increase the social expenditure in the short term, and this was the case in our experience. However, nursing home costs typically exceeds domiciliary care costs (nearly four times in our area) and have been identified among long-term services with higher contribution to the economic burden of stroke [18]. Therefore, in the mid- and long-term, early delivery of domiciliary care in stroke patients would alleviate the financial burden of the whole system. This effect, however, would not manifest unless pooling budget policies, based on a joint health and social care funding, are implemented.

One of the key elements of an integrated care program for early delivery of both social and health care services is the adequate integration between the two information systems, which in our country ―as well as in most countries― are handled by different entities and often have non-overlapping access profiles. In our experience, the integration of these two information systems, in force by the time of designing the RHP program, was a mainstay for a successful deployment of the integrated care approach. One of the most relevant consequences of the integration of the two information systems includes avoiding redundant assessments by the health and social care teams, which in our area ―like in most countries― belong to different government departments and entities, responsible to provide these services. In this regard, a systematic integration of health and social information systems would ease benchmarking analyses as well as health and social care planning.

Our study is strengthened by the inclusion of a population-based control group, which was feasible thanks to the central storage of healthcare data from nearly the entire population of Catalonia, as described elsewhere [9,10]. On the other hand, pairing of individuals in the RHP program and those in the control group was limited to the variables collected in central records. These records do not contain data related to the stroke episode such as stroke severity according to the National Institute of Health Stroke Scale, and relevant patient information such as the Barthel score for physical disability or the Pfeiffer scale for cognitive impairment. Of note, although stroke severity could not be included in the pairing, recipients of the RHP program were selected according to their health and social needs and are, therefore, expected to have poorer health status than those in the control group. Finally, the retrospective nature of the analysis precluded the administration of validated questionnaires for assessing the quality of care.

In summary, our analysis showed that an integrated care program of health and social care for stroke patients at discharge successfully promotes early domiciliary care delivery, resulting in a less frequently and later institutionalization in a long-term care facility. The integration of health and social care information systems enables not only the appropriate coordination of the various stakeholders, but also the analysis of the outcomes of care delivery. The benefits of post-discharge integrated care services for stroke patients are likely to increase in the context of pooling budget approaches that consider and plan social and health care services as a whole.

## Data Availability

All data produced in the present study are available upon reasonable request to the authors

## Ethics statement

The Independent Ethics Committees of each of the participating hospitals approved the study protocol. Participants provided informed consent.

