## Supplementary file 1 for "Data and care integration for post-acute intensive care program of stroke patients: effectiveness assessment using a disease-matched comparator cohort"

Supplementary appendix

**Figure S1.** Flow-chart of the integrated care intervention

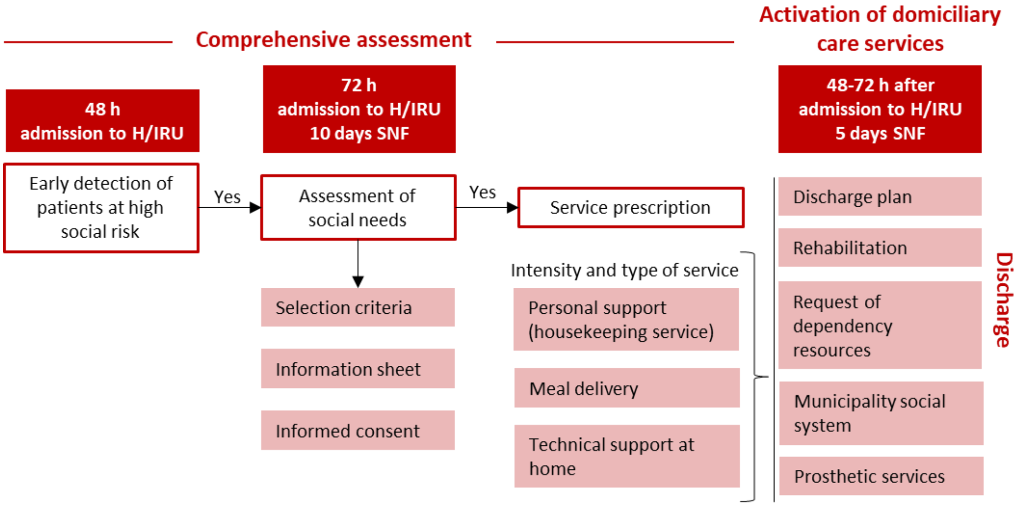


**H:** Hospital. **IRU:** Intensive rehabilitation unit. **SNF:** skilled nursing facility

**Table S1.** Social and health care expenditure after discharge

| Period | Group | Health care expenditure | | | | | Social care expenditure | Total expenditure |
|---|---|---|---|---|---|---|---|---|
| | | Outpatient resources | Inpatient resources | Pharmacy | Other | **Total Healthcare** | | |
| **Months 1-12** | **Control** | 117.48 | 398.54 | 110.46 | 29.06 | 655.54 | 145.63 | **801.17** |
| | **Intervention** | 132.94 | 425.88 | 96.19 | 81.00 | 736.01 | 251.01 | **987.02** |
| **Months 2-12** | **Control** | 112.49 | 248.65 | 110.04 | 29.56 | 500.75 | 156.40 | **657.15** |
| | **Intervention** | 133.10 | 211.90 | 97.87 | 83.77 | 526.64 | 257.95 | **784.58** |

**Figure S2.** Monthly use of resources within the 24 months following discharge in the integrated care (intervention) group (**A**) and population-based control group (**B**).

#### Domiciliary rehabilitation

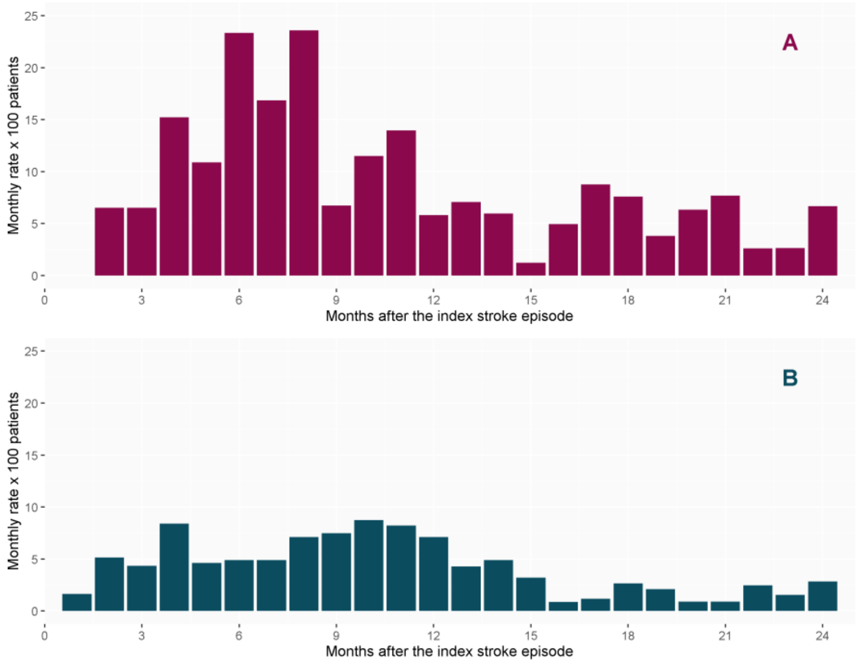


#### Emergency hospital admissions

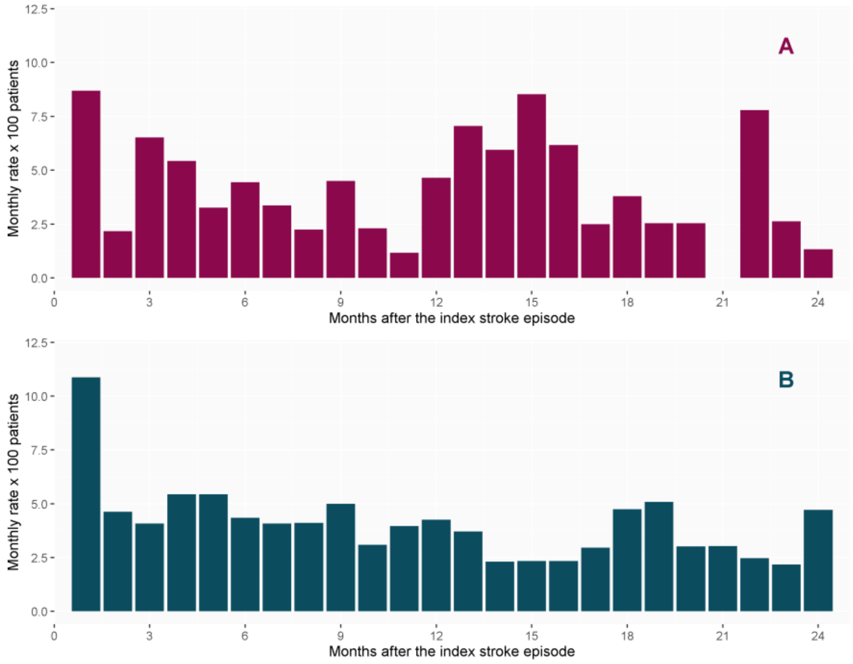

### Admissions to the emergency department

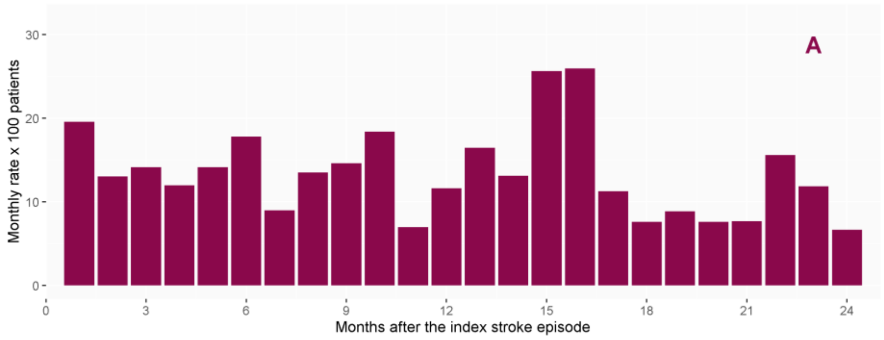


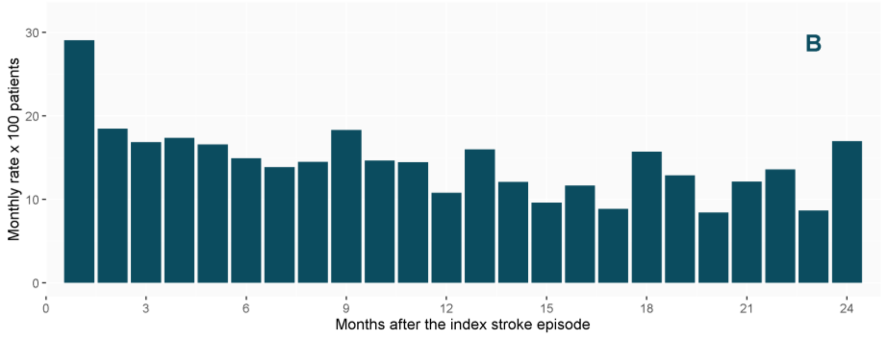


### Non-emergency medical transport rides

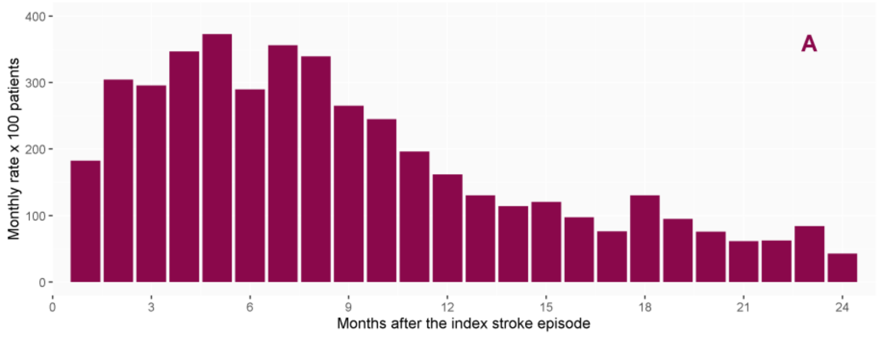


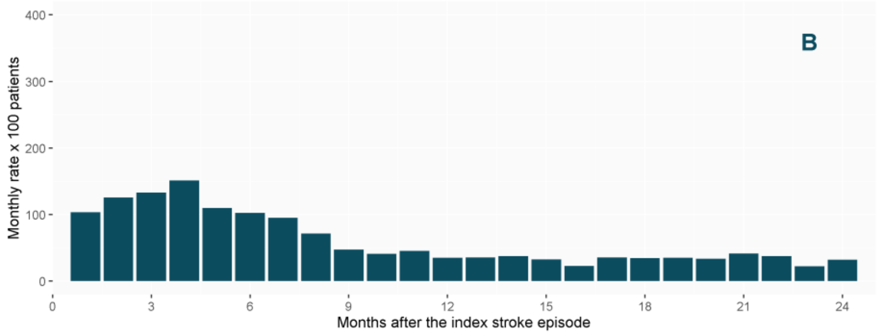

### Visits to primary care nurse staff

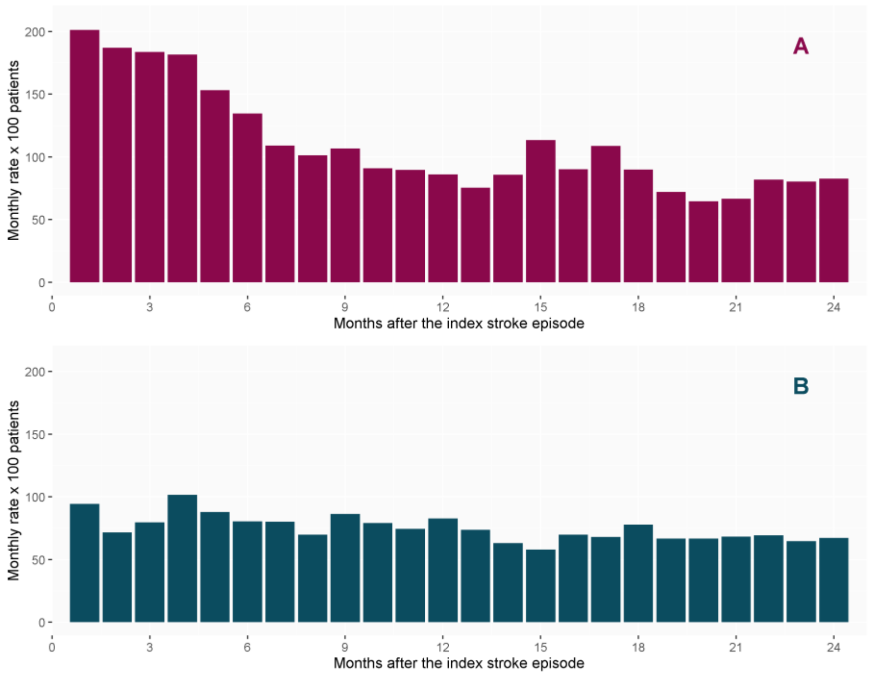


### Visits to primary care social worker

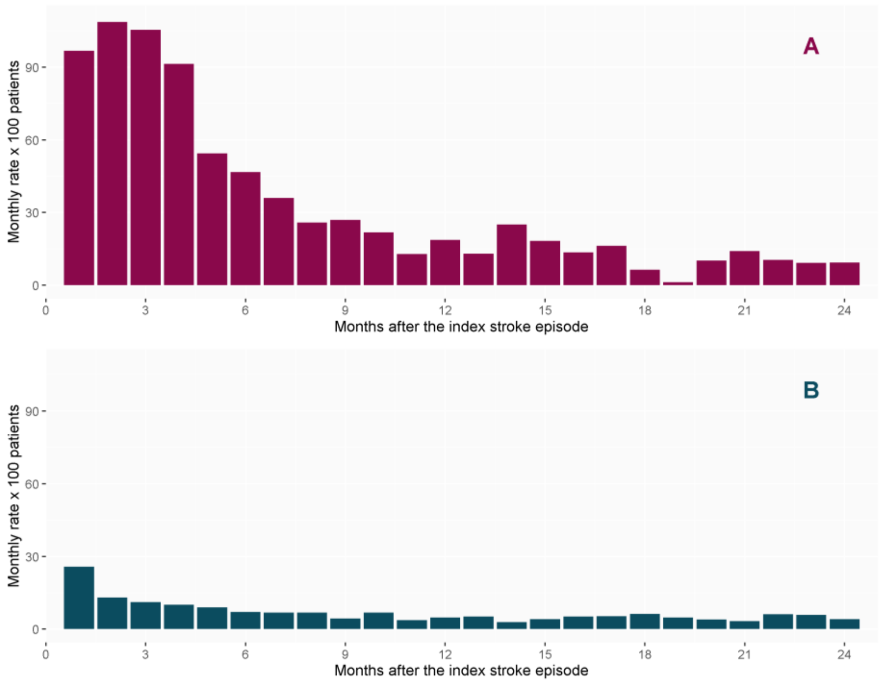

### Hospital admissions

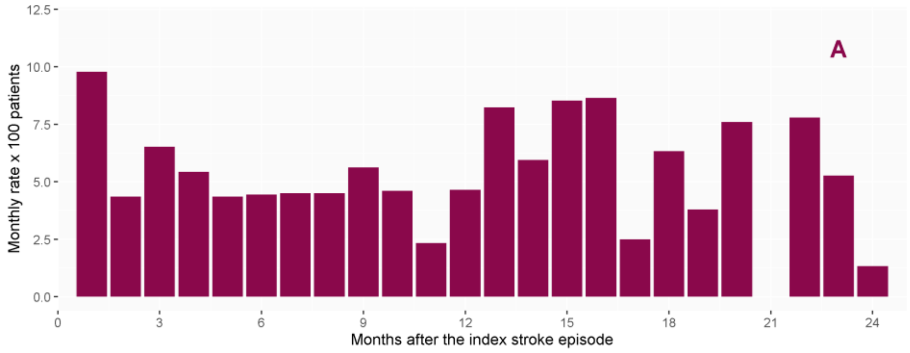


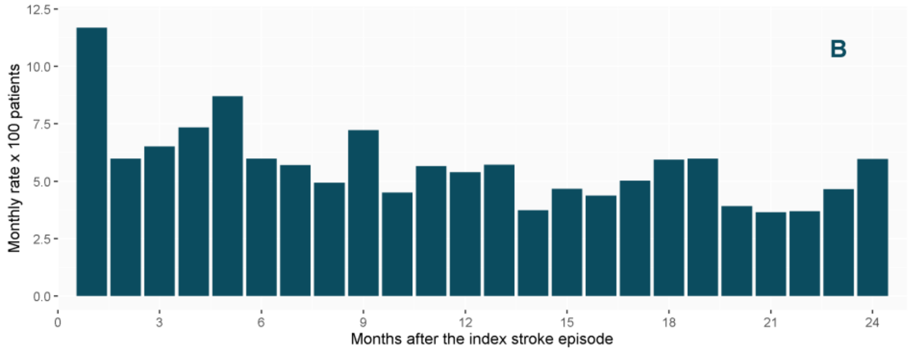


### Primary care visits

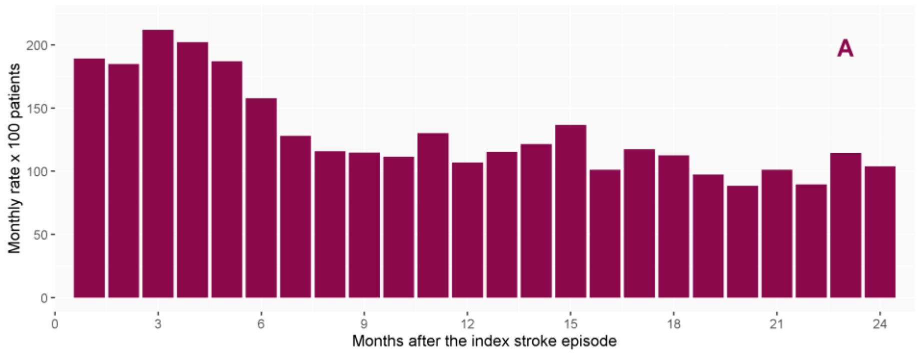


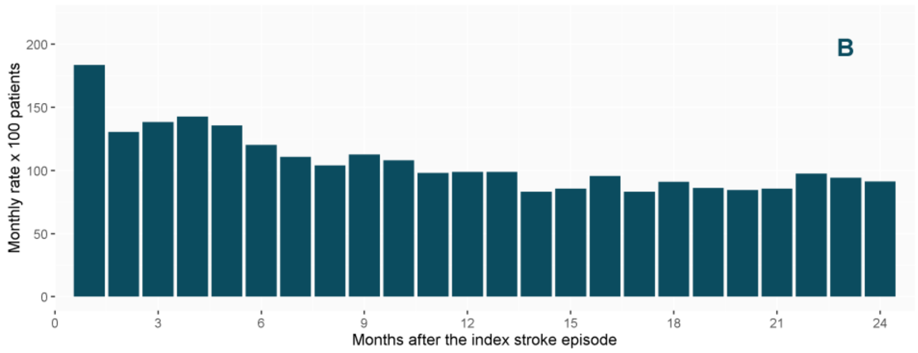

**Visits to the general practitioner**

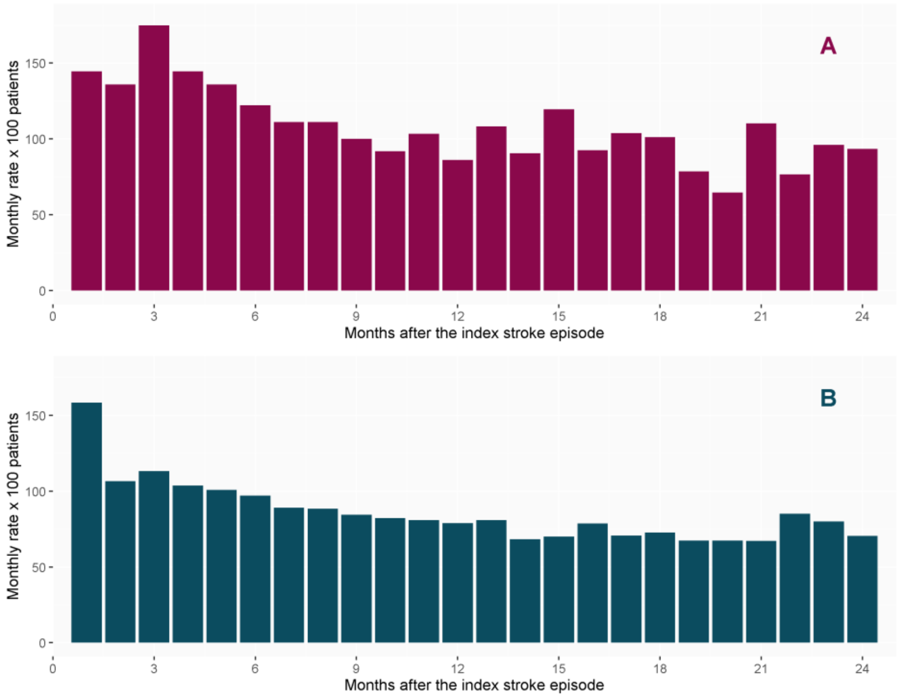


**Pharmacy (dispensed packages)**

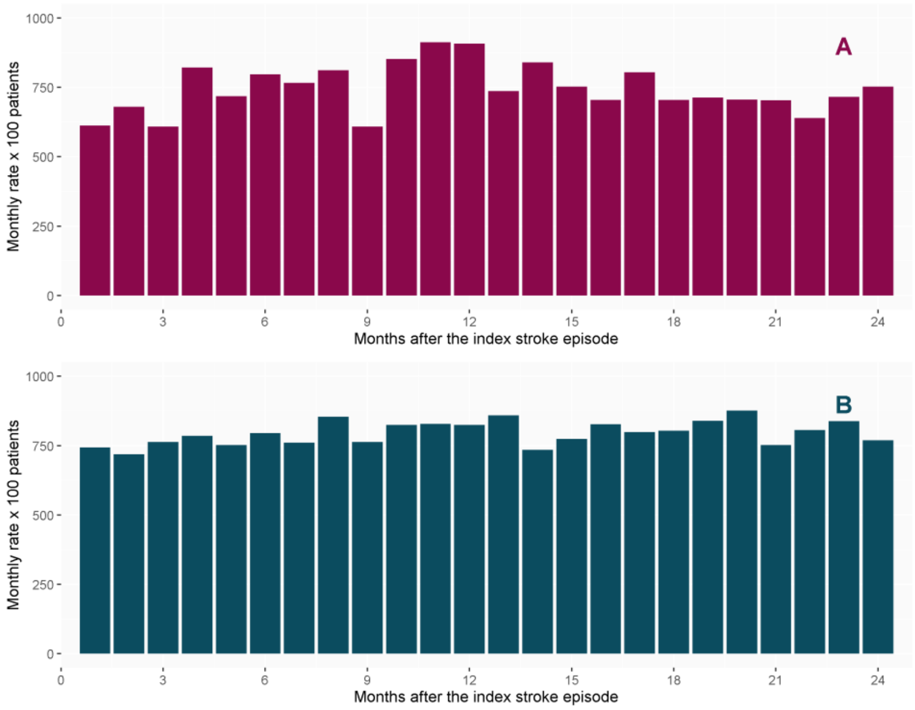

### Remote consultations

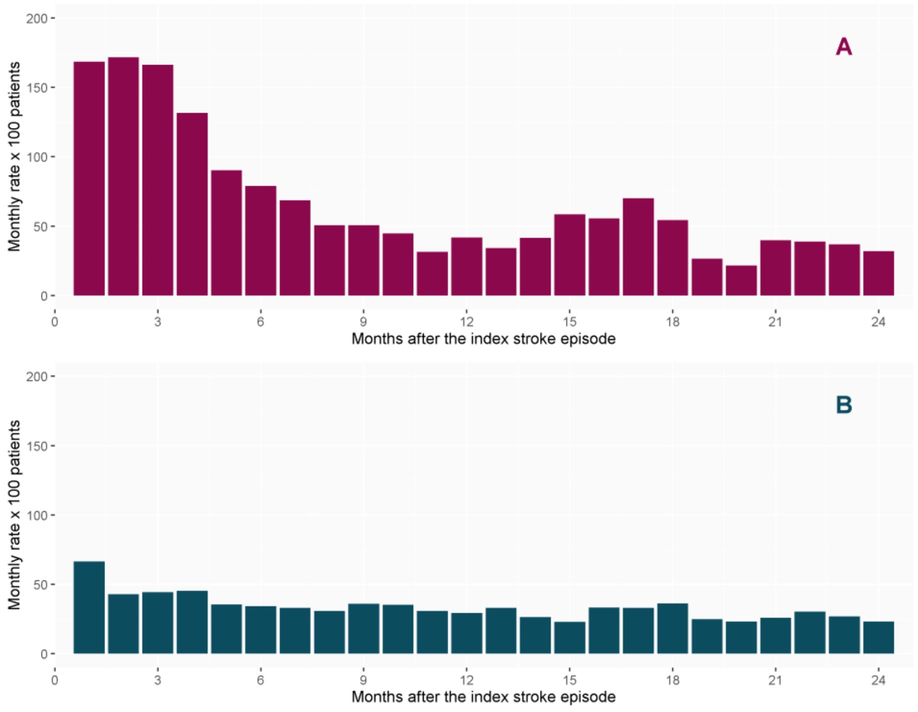


### Visits to the primary care center

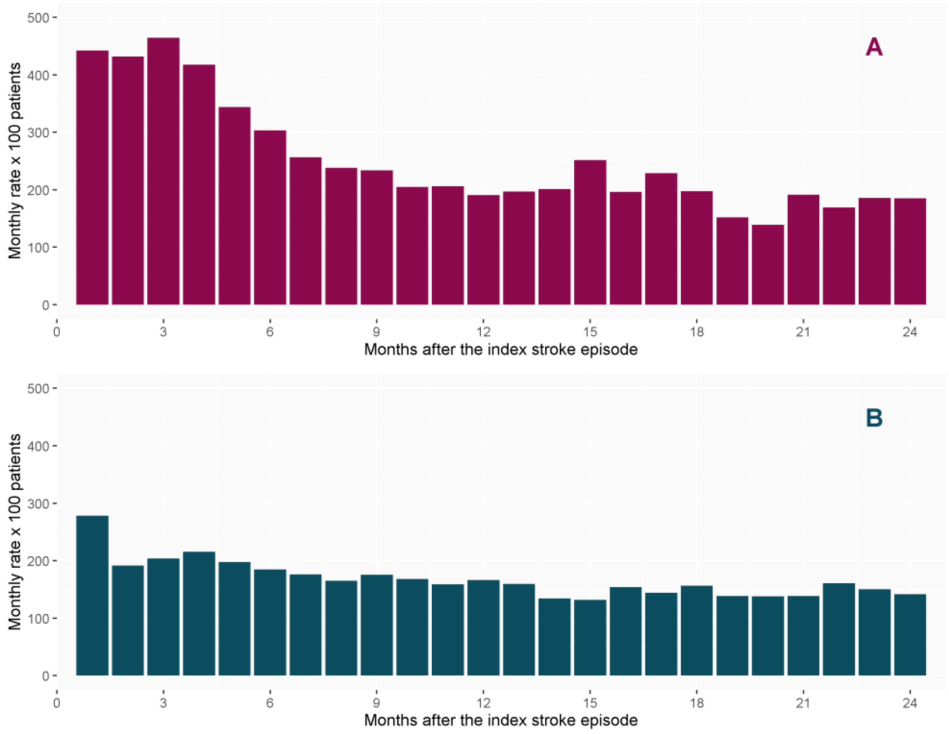

### Home care (primary care team) visits

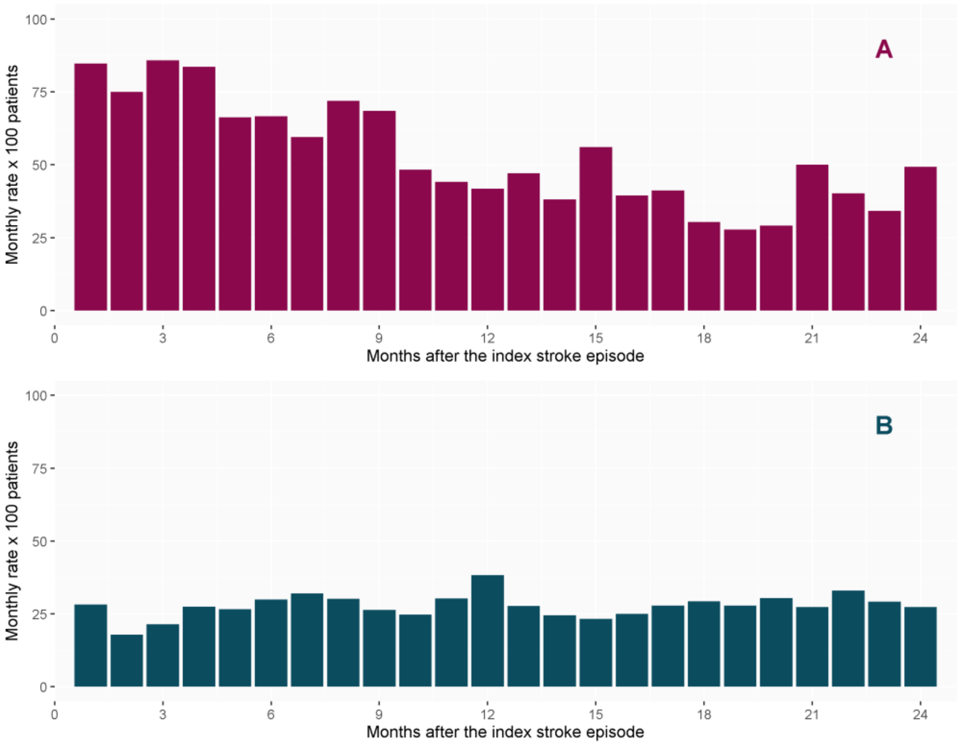


### Outpatient visits

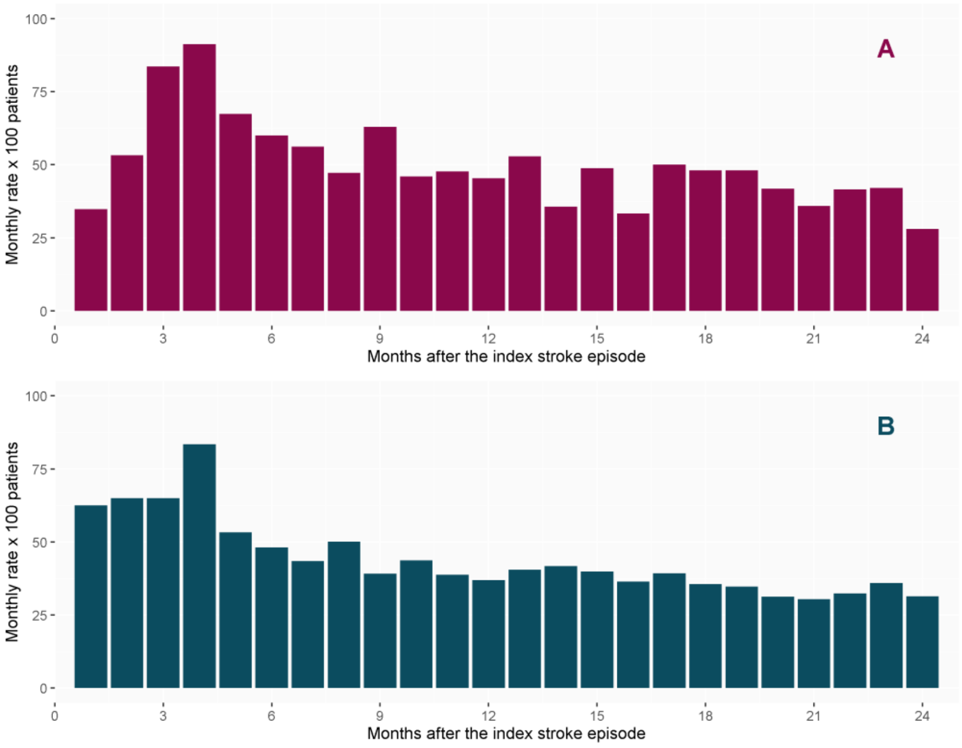
